# ANALYSIS OF SELF-CARE FACTORS IN HYPERTENSION PATIENTS IN COMMUNITY SETTINGS

**DOI:** 10.1101/2024.12.01.24318276

**Authors:** Ni Putu Ayu Ratna Dewi, Moses Glorino Rumambo Pandin, Nursalam, Ni Luh Putu Inca Buntari Agustini, Ni Wayan Sri Wahyuni, Komang Ardidhana Nugraha Putra

## Abstract

**Background:** Controlling blood pressure in hypertensive patients is one of the most important interventions in preventing complications, reducing morbidity, and premature mortality. Patients must practice hypertension self-care to prevent the disease’s frequent recurrence from deteriorating their health and to maintain effective behavior.

**Objective:** Analyze the factors that influence the implementation of self-care in hypertensive patients.

**Method:** Cross-sectional using the purposive sampling technique involving 120 respondents. The study was conducted in the working area of the Gianyar 1 Health Center, Bali, Indonesia. Data collection utilized the Indonesian version of the High Blood Pressure self-care profile questionnaire. Analysis included univariate, bivariate using chi-square, and multivariate using multinomial logistic regression.

**Results:** A long history of suffering from hypertension has an effect on behavior; education has an effect on motivation; and education and work have an effect on self-efficacy.

**Conclusion:** The article emphasizes the importance of holistic treatments in managing high blood pressure, with a focus on patient engagement to improve taking care of themselves with focused education and support systems. The outcomes align with the axiological dimension of philosophy by highlighting practical values that enhance patients’ autonomy and quality of life through education and supportive systems. Further, the research makes an empirical contribution by providing a comprehensive knowledge of self-care characteristics, which can be used to develop culturally appropriate and patient-focused intervention techniques.

## BACKGROUND

Hypertension, as a chronic illness, demands a complex perspective that incorporates biological, social, and philosophical dimensions. The fundamental idea of self-care is in line with axiological ideas, emphasizing patient autonomy and wellbeing. In 2023, hypertension claimed the lives of 10.8 million individuals, while 46% of the 1.28 billion people who had the condition were not aware they had it (World Health Organization, 2023b). ver the last four decades, the prevalence of hypertension has migrated from developed to low- and middle-income countries around the world. In underdeveloped nations like Indonesia, over three-quarters of hypertension sufferers reside (Konlan & Shin, 2023). The estimated prevalence of hypertension in Indonesia was 25.8% in 2013, and it rose to 34.1% in 2018 (Indonesian Ministry of Health, 2018). Specifically, in Bali Province, cases of hypertension in 2018 reached 30% (Central Bureau of Statistics, 2021) while in 2021 it reached 555,184 people (Bali Provincial Health Service, 2022).

Hypertension is mostly found in the elderly, but it is undeniable that hypertension can occur in all age groups (Winarto et al., 2021). Research by Fryar *et al*. (2019) in the United States stated the prevalence of hypertension in adults aged 18-39 years (7.5%), aged 40-59 years (33.2%), and aged 60 years and over (63.1%). In addition to age, the incidence of hypertension is influenced by various other factors such as genetics, sodium consumption, smoking, poor sleep quality, excessive alcohol consumption, and stress (Jung & Moon, 2023). Hypertension as a chronic disease, if not treated and controlled, will cause complications such as stroke, heart failure, kidney failure, cerebrovascular disease, and even death. Controlling blood pressure in hypertensive patients is one of the most important interventions in efforts to prevent complications, reduce morbidity, and premature mortality. Lorber & Divjak (2022) also stated that a reduction in blood pressure of 10 mmHg can reduce heart failure by 28%, stroke by 27%, coronary heart disease by 17%, and overall mortality by 13%.

The perspective of philosophy considers self-care for high blood pressure as greater than just a set of behaviors, it’s a complex phenomenon influenced by social, cultural, and individual factors (Alzahrani et al., 2022; Nazeri et al., 2022; Schaefer et al., 2022). Patients’ perceptions of their health and ability to manage their illness are influenced by their interactions with their environment, including cultural values, healthcare accessibility, and family (Leonardo et al., 2023; Pahria et al., 2022). Regarding hypertension, the Health and Social Security Administering Agency (BPJS Kesehatan) has created the Prolanis Chronic Disease Management Program. Regulation Number 7 of 2019 explains that the Chronic Disease Management Program (Prolanis) is a health service system through a proactive approach that is implemented in an integrated manner involving participants, health facilities, and the Health and Social Security Administering Agency in order to maintain health for participants suffering from chronic diseases to achieve optimal quality of life with effective and efficient health service costs. However, the level of participation is still not optimal, Khoe *et al*. (2020) stated that only 0.02% of the high-risk population for hypertension utilized Prolanis, so these efforts have not had an optimal impact.

WHO states that only about one in five hypertension sufferers are able to control their disease (World Health Organization, 2023a). Indonesian basic health research (2018) states that in Indonesia only 8.8% of hypertension patients routinely undergo treatment. Factors that influence this phenomenon are: use of the wrong dose and/or inadequate combination of drugs, poor adherence to treatment, self-perception of feeling healthy, unhealthy lifestyle (smoking, alcohol abuse, excess fat and salt, lack of activity, and overweight) (Villafuerte *et al*., 2020). This highlights the axiological importance of approaching self-care not only as a medical treatment but also as a complete strategy for reducing the global health burden (Gelaw et al., 2021). This study enhances quality of life by allowing patients to have more control over their medical condition while simultaneously cutting healthcare costs and system redundancy.

The existing hypertension control program’s shortcomings necessitate the development of a more comprehensive and innovative strategy among stakeholders. The newly developed intervention program must target more than simply the individual (beyond the individual), given that individuals cannot be divorced from their surroundings and social context (Fauzi *et al*., 2020). Calisane *et al*. (2021) believe that self-care is a crucial component of treatment that empowers individuals. In general, increasing patient awareness of the need for self-care might help patients take an active role in their disease management (Or *et al*., 2020). Even though self-care is essential to the effective management of hypertensive patients, there is a dearth of studies in nursing regarding such patients, particularly when related to using a self-care strategy. Furthermore, the current research site remains focused on the Denpasar and Badung districts, Bali, Indonesia, despite the fact that the Bali Provincial Health Profile data (2022) shows Gianyar Regency as having the second highest number of hypertension patients (Dinas Kesehatan Provinsi Bali, 2022).

The goal of this study is to provide a preliminary overview of the care of hypertension in Gianyar Regency, Bali, Indonesia. Through an epistemological lens, the study aims to generate actionable insights that inform patient-centered and culturally relevant interventions. It is hoped that these findings will not only advance theoretical understanding but also provide practical solutions to improve the quality of life for hypertensive patients.

## METHOD

A descriptive method was used in the study, which had a cross-sectional design. The cross-sectional technique was chosen because it provides a comprehensive snapshot of self-care features, capturing relationships across variables at a single point in time (Nursalam, 2020). This technique is in line with the epistemology goal of investigating how knowledge about self-care is produced, measured, and implemented in order to produce practical findings that can guide future initiatives. The study was carried out between August and September 2024 in the working area of the Gianyar 1 Health Center, Bali, Indonesia, which includes six villages and four sub-districts.

The Slovin formula was used to calculate the sample size of 109 respondents. To account for dropouts, the projected sample size was adjusted by 10%, resulting in a total of 120 respondents for this study. The sampling method utilized was deliberate sampling. Inclusion criteria: aged 40– 60 years, diagnosed with primary hypertension within the last year and following medication, and able to converse verbally in Indonesian or Balinese that the researcher understands. Exclusion criteria: suffering from critical hypertension with systolic ≥ 180 mmHg and diastolic ≥ 120 mmHg or who have other comorbidities.

The researcher has applied to the Research Ethics Commission of ITEKES Bali, number: 03.0233/KEPITEKES-BALI/VII/2024. The study’s tools included a questionnaire on demographics and a high blood pressure self-care profile (HBP-SCP) questionnaire created by Han *et al*. (2014). The HBP-SCP questionnaire contains three subscales: behavior, motivation, and self-efficacy. The Indonesian version of the HBP-SCP questionnaire was translated and validated by Upoyo *et al*. (2021), and the Cronbach’s alpha value was greater than 0.70.

The tool’s emphasis on the shifting and complicated nature of personal hygiene coincides with the ontological view that a range of systemic and personal elements influence self-care behaviors. This study employs univariate analysis (frequency distribution tables and percentages), bivariate analysis (chi-square), and multivariate analysis (multinomial logistic regression). These approaches were utilized to identify critical relationships and create the framework for evidence-based interventions to improve self-care in hypertension patients.

## RESULT

According to 120 respondents, the highest age range is 61-70 years, with 36 (30%). In terms of gender, women account for the majority of responses, 64 (53.3%). The majority of respondents (28.3%) have a high school education. Due to the fact that the majority of people in the 61–70 age group are considered elderly, not working receives the highest ranking of 39 (32.5%). The majority of respondents have had hypertension for 1–5 years, 65 (54.2%).

Based on table 2, it shows that the majority of respondents’ self-care behavior is moderate, as many as 81 (67.5%), the majority of respondents’ self-care motivation is in the moderate category, 88 (73.3%), and the respondents’ self-care self-efficacy is in the moderate category, as many as 73 (60.8%).

**Table 1.** Frequency distribution of respondent characteristics (n=120)

| Characteristics | Frequency (f) | Percentage (%) |
| --- | --- | --- |
| **Age (year)** |  |  |
| 41-50 | 25 | 20.8 |
| 51-60 | 34 | 28.3 |
| 61-70 | 36 | 30.0 |
| 71-80 | 25 | 20.8 |
| **Mean** |  | 61.11 |
| **SD** |  | 9.539 |
| **Sex** |  |  |
| Male | 56 | 46.7 |
| Female | 64 | 53.3 |
| **Religion** |  |  |
| Hinduism | 94 | 78.3 |
| Islam | 16 | 13.3 |
| Christianity | 10 | 8.3 |
| **Education** |  |  |
| No school | 19 | 15.8 |
| Elementary school | 22 | 18.3 |
| Junior high school | 30 | 25.0 |
| Senior high school | 34 | 28.3 |
| Bachelor's Degree/Diploma | 15 | 12.5 |
| **Occupation** |  |  |
| Doesn't work | 39 | 32.5 |
| Private | 23 | 19.2 |
| Housewife | 20 | 16.7 |
| Civil servants | 3 | 2.5 |
| Farmer | 9 | 7.5 |
| Other | 26 | 21.7 |
| **Duration of hypertension (year)** |  |  |
| 1-5 | 65 | 54.2 |
| 6-10 | 52 | 43.3 |
| 11-15 | 3 | 2.5 |

**Table 2.** Implementation of self-care that has been carried out by hypertension patients.

| Characteristics | f | % | Mean | SD |
| --- | --- | --- | --- | --- |
| **Behavior** |  |  |  |  |
| Good | 20 | 16.7 |  |  |
| Moderate | 81 | 67.5 | 46.04 | 8.686 |
| Bad | 19 | 15.8 |  |  |
| **Motivation** |  |  |  |  |
| Poor | 17 | 14.2 |  |  |
| Moderate | 88 | 73.3 | 53.04 | 12.432 |
| High | 15 | 12.5 |  |  |
| **Self-Efficacy** |  |  |  |  |
| Poor | 25 | 20.8 |  |  |
| Moderate | 73 | 60.8 | 48.20 | 8.776 |
| High | 22 | 18.3 |  |  |

Based on table 3, there is no statistically significant link between age, gender, occupation, and education and self-care behavior (p-values 0.717, 0.688, 0.270, and 0.602). While the duration of hypertension is significantly associated with individual self-care behavior (p-value = 0.048), There is no association between age, gender, and duration of hypertension and self-care motivation (p-values of 0.607, 0.857, and 0.232, respectively). Individual self-care motivation is substantially correlated with occupation and education (p-value = 0.002 and <0.001, respectively). Meanwhile, efficacy analysis reveals that age, gender, and duration of hypertension had no link with self-care self-efficacy (p-value = 0.506; 0.161; 0.204). Meanwhile, work and education are significantly related to individual self-care self-efficacy with a p-value of 0.010 and <0.001.

**Table 3.** Relationship between respondent characteristics with behavior, motivation, and self-care efficacy (n = 120)

| Variable | Behavior |  | Motivation |  | Self-Efficacy |  |
| --- | --- | --- | --- | --- | --- | --- |
|  | X<sup>2</sup> | P-value | X<sup>2</sup> | P-value | X<sup>2</sup> | P-value |
| Age | 3.705 | 0.717 | 4.516 | 0.607 | 5.303 | 0.506 |
| Sex | 0.749 | 0.688 | 0.309 | 0.857 | 3.652 | 0.161 |
| Occupation | 12.232 | 0.270 | 27.495 | 0.002 | 23.164 | 0.010 |
| Education | 6.401 | 0.602 | 95.745 | <0.001 | 29.031 | <0.001 |
| Duration of hypertension | 9.567 | 0.048 | 5.586 | 0.232 | 5.935 | 0.204 |

\*Chi-Square

Based on table 4, it shows that the behavioral variables: age, gender, occupation, and education do not affect individual self-care behavior. However, a long history of suffering from hypertension affects self-care behavior. Motivational variables: age, gender, duration of hypertension, and occupation do not affect individual self-care motivation. However, education affects self-care motivation. While the efficacy variable shows that age, gender, and duration of hypertension do not affect individual self-care efficacy. However, education and occupation affect self-care self-efficacy. These results indicate the possibility for focused educational and occupational treatments to enhance hypertension patients’ self-care routines.

**Table 4.** Analysis of respondent characteristics with behavior, motivation, and self-efficacy (n = 120)

| Variable | Behavior |  | Motivation |  | Self-Efficacy |  |
| --- | --- | --- | --- | --- | --- | --- |
|  | Sig. | Nagelkerke R Square | Sig. | Nagelkerke R Square | Sig. | Nagelkerke R Square |
| Age | 0.243 |  | 0.479 |  | 0.265 |  |
| Sex | 0.383 |  | 0.486 |  | 0.792 |  |
| Occupation | 0.710 | 35.7% | 0.092 | 74.6% | 0.004 | 48.4% |
| Education | 0.064 |  | <0.001 |  | 0.008 |  |
| Duration of hypertension | 0.003 |  | 0.901 |  | 0.473 |  |

## DISCUSSION

Self-care, as a cornerstone of chronic disease management, necessitates the development of specialized prevention, management, and therapy skills, which directly contribute to better health outcomes and lower healthcare costs (Hani et al., 2024; Putri et al., 2021; Sarfika et al., 2023). Prior study has examined self-care. in patients with chronic diseases that include those with diabetes, chronic obstructive pulmonary disease (COPD), heart failure, and hypertension, and found that developing the competencies required for handling these conditions led to more effective outcomes and controlling the disease (Villafuerte et al., 2020). It is essential to develop new and effective interventions for patients with uncontrolled hypertension, as these individuals have higher morbidity and mortality rates than patients with controlled hypertension. Based on the results of the study that has been conducted, it was found that for hypertensive patients in the Gianyar, Bali, Indonesia most of their self-care behavior, motivation, and self-efficacy were in the moderate category.

This study found that a long history of hypertension has a significant influence on self-care behavior. Individuals who have had hypertension for a long time tend to have a higher awareness of the risk of complications that can occur if the condition is not managed properly (Pahria et al., 2022). This awareness often encourages them to be more disciplined in carrying out self-care behaviors. Such findings highlight the potential of targeted interventions, particularly for patients with a long history of hypertension, to further enhance their self-care practices. Direct experience with chronic conditions such as hypertension often teaches individuals about the importance of self-management, including routinely monitoring blood pressure, following medication, and adopting a healthy lifestyle (Putri et al., 2021). This experience strengthens their commitment to self-care practices. Fear of complications such as heart attack or stroke may be the main driver for individuals with a long history of hypertension to be more serious about maintaining their health through self-care behaviors (Boitchi et al., 2021; Yeom, 2021).

This study emphasizes the practical value of education in enhancing self-care motivation and self-efficacy. Educational interventions that provide patients with knowledge and understanding about hypertension management can improve motivation and encourage proactive health behaviors (Schaefer et al., 2022). Education merely encourages motivation and self-efficacy, additionally represents the axiological element of philosophy, as it helps patients to make educated decisions and strengthen their autonomy to manage chronic diseases. Individuals with higher levels education might become better equipped to weigh the health benefits and risks of self-care, motivating them to take action (Kurt & Gurdogan, 2022). These treatments are consistent with global health priorities, reducing the burden of long-term illnesses while enabling people to take care of their own health. For example, promotional efforts highlighting the importance of regular medication consumption, regular BP checks, and lifestyle changes could be critical in improving patient outcomes (Howren et al., 2020). Furthermore, these interventions contribute to the larger goal of improving the quality of life for those with long-term conditions like hypertension.

These results support the epistemological theory that self-care is a learned behavior shaped by a person’s experiences as well as outside factors like social settings and education. By supporting the idea that persons with chronic illnesses have a better understanding of their health over time, which enhances their ability to manage their sickness, this research contributes to the body of knowledge (Pahria et al., 2022). This is consistent with self-efficacy theory, which emphasizes that belief in one’s ability to perform health behaviors is critical to successful self-care (Osokpo et al., 2021). For example, patients who receive structured education programs that include role-playing scenarios or peer group discussions are more likely to build confidence in their ability to adhere to medication regimens and engage in physical activity (Kurt & Gurdogan, 2022; Setiadi et al., 2022). In this study, education not only influenced motivation but also strengthened self-efficacy, suggesting that empowering patients with knowledge was a key factor in increasing their confidence in managing hypertension.

This study also aligns with the ontological aspect of philosophy that self-care is a dynamic phenomenon influenced by personal experiences, social context, and cultural influences. Individual hypertension experiences, as well as the social and cultural context in which patients reside, all have a significant impact on their self-care behaviors (Afik & Fikriana, 2021; Gelaw et al., 2021). For instance, in Gianyar, Bali, traditional healing practices and community-based support systems are integral to the healthcare experience. Social status, cultural standards, and availability of medical facilities may all have an impact on a person’s capacity to follow food guidelines or exercise regularly (Boitchi et al., 2021; Nazeri et al., 2022). Addressing hypertension self-care requires a holistic approach that integrates education, cultural context, and policy support (Sarfika et al., 2023). Local health centers could organize monthly workshops where patients learn practical self-care skills, such as managing their diet or monitoring blood pressure, while also addressing cultural misconceptions about hypertension (Yeom, 2021). These findings lay a foundation for future interventions aimed at empowering patients and fostering sustainable health behaviors. These results suggest that understanding self-care effectively requires taking cultural wellness and the immediate environment into consideration.

## CONCLUSION

Hypertensive patients in Gianyar, Bali, exhibit moderate levels of self-care behavior, motivation, and self-efficacy, with education and employment identified as key factors influencing these outcomes. This study highlights the need for comprehensive interventions that integrate pharmacological approaches, such as self-monitoring, with non-pharmacological strategies like lifestyle modifications tailored to local cultural contexts. These findings highlight the critical role of structured education and support networks in increasing patient autonomy, lowering the burden of hypertension, and improved quality of life. Furthermore, the ontological perception of self-care as an evolving relationship between personal and social elements emphasizes the necessity for comprehensive, culturally appropriate interventions. By recognizing self-care as a multidimensional phenomenon shaped by social, cultural, and systemic influences, this study provides a foundation for holistic, patient-centered hypertension management models and underscores the need for culturally relevant health policies

## IMPLICATION

These data support the notion that self-care is a fluid construct impacted by social, cultural, and environmental factors. The significance of investigating non-biomedical elements, such as customs and community dynamics, within a chronic disease treatment framework is highlighted by incorporating cultural context into self-care management. This study bridges empirical findings with philosophical dimensions by demonstrating the axiological importance of interventions that enhance patient autonomy and quality of life. It also contributes to the epistemological understanding of self-care as a learned behavior shaped by experience and education, while recognizing the ontological significance of cultural and social contexts in healthcare. The implementation and acceptance of self-care treatments may be improved by utilizing Gianyar, Bali’s pre-existing community structures, such as traditional support networks and community health facilities. In order to promote better adherence to self-care behaviors, health providers must create programs that include regional cultural customs, such as patient-centered interventions.

## LIMITATION

This study has been conducted as optimally as possible, but there are several limitations of this study, namely: 1) This study is a cross-sectional design, so there are limitations in concluding causal relationships between variables. 2) This study was conducted in a specific area (Gianyar, Bali), so that the generalization of the research results is limited to other populations or areas that have different cultural or socio-economic contexts. 3) This study only looks for factors that influence self-care and does not explore in depth certain cultural practices or beliefs that can influence behavior. Further research can investigate how cultural aspects influence self-care routines and how to incorporate them into health interventions. Research on the efficiency of culturally tailored self-care programs can provide insight into how to improve chronic disease management, especially in diverse groups.

## Data Availability

All data produced in the present study are available upon reasonable request to the authors
All data produced in the present work are contained in the manuscript

## ACKNOWLEGMENTS

Thank you to the nursing lecturers at Airlangga University and very grateful that the Faculty of Nursing at Airlangga University has provided facilities for this research

